# Sexual violence in older adults: a Belgian prevalence study

**DOI:** 10.1101/2021.03.04.21252934

**Authors:** Anne Nobels, Adina Cismaru Inescu, Laurent Nisen, Bastien Hahaut, Marie Beaulieu, Gilbert Lemmens, Stéphane Adam, Evelyn Schapansky, Christophe Vandeviver, Ines Keygnaert

**Author notes:** Direct correspondence to: Anne Nobels, International Centre for Reproductive Health, Ghent University, C. Heymanslaan 10 entrance 75- ICRH, 9000 Gent, Belgium.

## Abstract

**Background:** Sexual violence (SV) is an important public health problem which may cause long-lasting health problems. SV in older adults remains neglected in research, policies and practices. Valid SV prevalence estimates and associated risk factors in older adults are currently unavailable.

**Objective:** To measure lifetime and past 12-months sexual victimisation in older adults living in Belgium, its correlates, assailant characteristics and the way that victims framed their SV experiences.

**Design:** Cross-sectional general population study.

**Setting:** Community-dwelling, assisted living and nursing homes.

**Participants:** 513 people of 70 years and older living in Belgium.

**Methods:** SV was measured using behaviourally specific questions based on a broad definition of SV. Participants were selected via a cluster random probability sampling with a random route finding approach. Information on sexual victimisation, correlates, assailant characteristics and framing was collected via structured face-to-face interviews.

**Results:** Lifetime SV prevalence was 44% (55% F, 29% M). Past 12-months prevalence was 8% (9% F, 8% M). Female sex and a higher number of sexual partners were associated with lifetime SV (p <.05), non-heterosexual sexual orientation with past 12-months SV (p <.05). Correlates generally linked to elder abuse and neglect were not linked with SV. ‘Someone unknown’ was identified as most common assailant.

**Conclusions:** SV appears to be common in older adults in Belgium. Both correlates and assailant characteristics seem to differ from previous studies on elder abuse and neglect. Recognising older adults as a risk group for sexual victimisation in research, policies and practices is of the utmost importance.

## 1. Introduction

Sexual violence (SV) [1] is increasingly considered a public health problem of major societal and judicial concern [2, 3]. Extensive research links sexual victimisation to long-lasting sexual, reproductive, physical, and mental health problems [2-4]. Exposure to childhood sexual abuse has been linked to depression, anxiety, and somatic complaints in older adults [5, 6].

Previous research suggests that SV in older adults rarely occurs [7]. A recent meta-analysis showed that 0.9% of community-dwelling older adults worldwide were sexually victimised in the past 12-months [8]. In Europe, numbers of past 12-months SV prevalence in older adults varied between 0% and 3.1% [9]. In a Belgian study, lifetime SV prevalence was estimated at 6.3% [10]. However, current studies show low SV prevalence numbers as they conflate it with other types of violence in the broader context of elder abuse and neglect [9], domestic violence or intimate partner violence [11]. Studies exclusively focussing on SV in older adults, describe criminal cases, and judicial response [12, 13]. Yet, research on SV in older adults from a public health perspective, providing valid SV prevalence numbers and correlates, is currently lacking.

Assessing sexual victimisation in older adults may be challenging for myriad reasons. Older adults grew up in a time when talking about sexuality and SV was considered taboo. They may also have different perceptions of sexuality and SV compared to younger generations [14], because of limited sexual education when they were young, different legal definitions and ideas on sexual consent [15, 16]. Furthermore, older adults are considered asexual by society [17]. Internalizing this stereotypical image of ‘the asexual older adult’, they may not identify themselves as possible SV victims [18, 19], which could lead to a reluctance to disclose sexual victimisation, and to seek help [11, 18]. Moreover, health care workers feel that sexuality and SV are not legitimate topics to discuss with older adults and are worried to offend their patients when they do so [20, 21]. Also, they seem to have insufficient communication skills to adequately deal with SV in later life [22].

In spite of the call by the United Nations to significantly reduce all forms of violence [23], policies on SV in older adults are currently non-existent [9]. In order to develop preventive measures and to provide tailored care for older SV victims, a revision of current policies and health care practices is of the utmost importance [9, 24]. To make this possible, a better understanding of the prevalence and nature of SV in older adults is crucial. To our knowledge, this study is the first in its kind to assess lifetime and past 12-months sexual victimisation, correlates, assailant characteristics and the way that older victims framed their SV experiences. Based on the results, we identify avenues for future research, and formulate recommendations for policies and health care practices.

## 2. Methods

### 2.1 Measures

We adopted the WHO definition of SV, which includes different forms of sexual harassment without physical contact, sexual abuse with physical contact but without penetration and (attempted) rape [1, 3]. This definition was expanded to include sexual neglect, as a result of recent insights in the field of SV in older adults [9, 25]. Participants were asked, among others, questions on sociodemographic characteristics, sexual health & relations and sexual victimisation. In order to provide valid estimates of both female and male sexual victimisation, we used behaviourally specific questions (BSQ) to assess lifetime and past 12-months SV experiences [26]. The SV items were based on existing surveys [27-29], and adapted to the Belgian social and legal context [30]. Due to the absence of a standardised measure for sexual neglect, it was assessed as “touching in care” (see Appendix 1).

### 2.2 Sample selection

Between the 8^th^ July 2019 and the 12^th^ March 2020, 513 older adults across Belgium were interviewed. Based on our power analysis, the target sample size was 845 participants [31]. It was anticipated this sample size would provide a SV prevalence estimate with a three percent margin of error. However, the COVID-19 pandemic and associated lockdown measures forced us to prematurely stop data collection. Cluster random probability sampling was used to obtain representative results for the Belgian older population. Eligible participants were identified using a random walk procedure [31, 32]. Participants had to be at least 70 years old, live in Belgium, speak Dutch, French or English, and have sufficient cognitive ability to complete the interview. Both older adults living in the community and living in nursing homes or assisted living facilities were included. Face-to-face interviews were carried out by trained interviewers in private at the participant’s place of residence.

The study was conducted according to the WHO ethical and safety recommendations for SV research [33] and received ethical approval from the ethical committee of Ghent University/University Hospital (B670201837542). All participants gave their informed consent before participating in the study. After participation they were given the contact details of several helplines. Participation rate was 34%. The full study protocol is available elsewhere [31].

### 2.3 Analyses

Statistical analysis was performed using R version 3.6.3 and SPSS Statistics 26. The 17 SV variables were grouped into hands-off (eight items) and hands-on SV (nine items), the latter being further grouped into sexual abuse (four items) and attempted or completed rape (five items). For the purpose of the analysis the item measuring sexual neglect was grouped under sexual abuse. We created dichotomous variables out of all items in order to assess lifetime and past 12-months victimisation. A detailed overview of the SV outcome measures can be found in Appendix 1.

A number of demographic and socio-economic variables and variables related to the participants’ sexuality were included in the multivariate logistic regression analysis. All variables were added simultaneously. Adjusted odds ratios describe the correlation with sexual victimisation while adjusting for the other variables in the model. The multi-collinearity assumption of multivariate regression analyses was tested with the Variance Inflation Factor (VIF) and indicated no violation. Social support (measured by number of confidants) could be added as a continuous variable into the model without violating the linearity assumption. The number of lifetime sexual partners and age of sexual initiation were recoded into dichotomous variables based on the median.

## 3. Results

### 3.1 Study population characteristics

The study sample consisted of a valid representation of the Belgian population aged 70 years and older [31]. The mean age was 79 years (SD: 6.4yrs, range 70-99yrs), 58.3% was female, 89.8% was community-dwelling, 90.4% was born in Belgium, 31.2% completed higher education, 50.3% was in a relationship and 7.4% labelled themselves as non-heterosexual. This group contains participants who labelled themselves as homosexual, bisexual, pansexual, asexual or other. In this last group, several participants labelled themselves as “normal”. Since it was not clear whether they had difficulties understanding the different terms defining sexual orientation or whether they indeed labelled their sexual orientation as “other”, we decided to classify these participants as non-heterosexual. More information on the sample composition can be found in Appendix 2.

### 3.2 Prevalence of sexual victimisation

The lifetime prevalence of SV was 44.2% (95% CI: 39.9-48.7), 55.2% (95% CI: 49.4-60.9) of females and 29.0% (95% CI: 23.0-35.5) of males. Almost half of women and one in four men experienced hands-off SV, one in three women and one in six men reported hands-on SV. One in twelve females and one in 30 males disclosed an (attempted) rape. In the past 12-months, 8.4% (95% CI: 6.1-11.1) experienced at least one form of SV, 7.0% (95% CI: 5.0-9.6) reported hands-off and 2.5% (95% CI: 1.4-4.3) hands-on SV. The most commonly reported sexually transgressive behaviours were unwanted sexual staring, sexual innuendo and kissing; both during lifetime and in the past 12-months. A more detailed description of the prevalence of all different forms of SV can be found in Table 1.

**Table 1.**
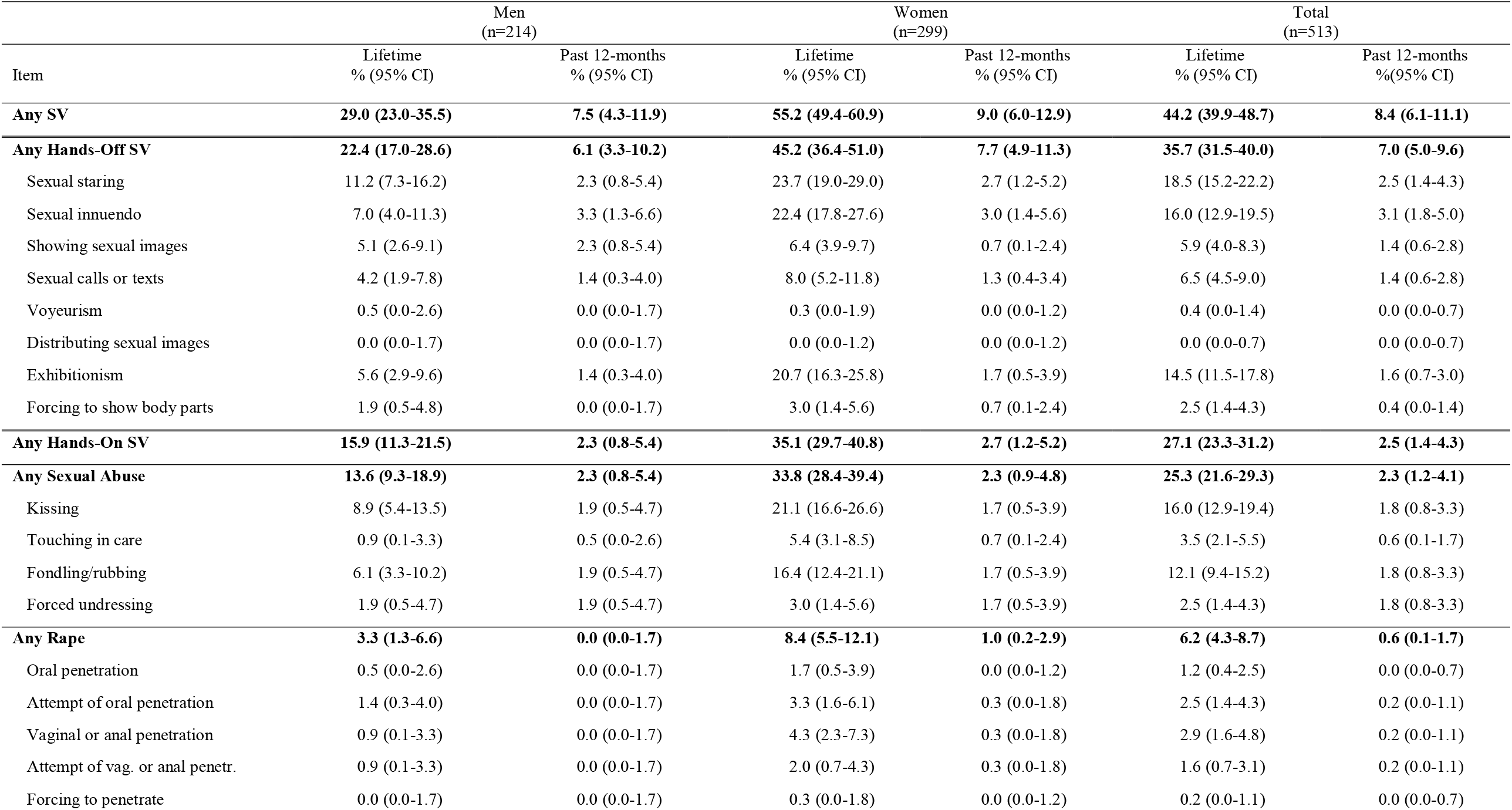
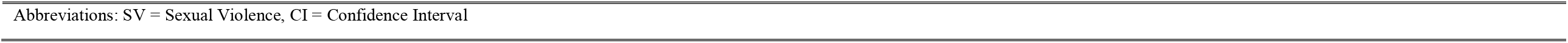
Detailed lifetime and past 12-months prevalence sexual victimisation, by sex

| Item | Men<br>(n=214) |  | Women<br>(n=299) |  | Total<br>(n=513) |  |
| --- | --- | --- | --- | --- | --- | --- |
|  | Lifetime<br>% (95% CI) | Past 12-months<br>% (95% CI) | Lifetime<br>% (95% CI) | Past 12-months<br>% (95% CI) | Lifetime<br>% (95% CI) | Past 12-months<br>% (95% CI) |
| **Any SV** | **29.0 (23.0-35.5)** | **7.5 (4.3-11.9)** | **55.2 (49.4-60.9)** | **9.0 (6.0-12.9)** | **44.2 (39.9-48.7)** | **8.4 (6.1-11.1)** |
| **Any Hands-Off SV** | **22.4 (17.0-28.6)** | **6.1 (3.3-10.2)** | **45.2 (36.4-51.0)** | **7.7 (4.9-11.3)** | **35.7 (31.5-40.0)** | **7.0 (5.0-9.6)** |
| Sexual staring | 11.2 (7.3-16.2) | 2.3 (0.8-5.4) | 23.7 (19.0-29.0) | 2.7 (1.2-5.2) | 18.5 (15.2-22.2) | 2.5 (1.4-4.3) |
| Sexual innuendo | 7.0 (4.0-11.3) | 3.3 (1.3-6.6) | 22.4 (17.8-27.6) | 3.0 (1.4-5.6) | 16.0 (12.9-19.5) | 3.1 (1.8-5.0) |
| Showing sexual images | 5.1 (2.6-9.1) | 2.3 (0.8-5.4) | 6.4 (3.9-9.7) | 0.7 (0.1-2.4) | 5.9 (4.0-8.3) | 1.4 (0.6-2.8) |
| Sexual calls or texts | 4.2 (1.9-7.8) | 1.4 (0.3-4.0) | 8.0 (5.2-11.8) | 1.3 (0.4-3.4) | 6.5 (4.5-9.0) | 1.4 (0.6-2.8) |
| Voyeurism | 0.5 (0.0-2.6) | 0.0 (0.0-1.7) | 0.3 (0.0-1.9) | 0.0 (0.0-1.2) | 0.4 (0.0-1.4) | 0.0 (0.0-0.7) |
| Distributing sexual images | 0.0 (0.0-1.7) | 0.0 (0.0-1.7) | 0.0 (0.0-1.2) | 0.0 (0.0-1.2) | 0.0 (0.0-0.7) | 0.0 (0.0-0.7) |
| Exhibitionism | 5.6 (2.9-9.6) | 1.4 (0.3-4.0) | 20.7 (16.3-25.8) | 1.7 (0.5-3.9) | 14.5 (11.5-17.8) | 1.6 (0.7-3.0) |
| Forcing to show body parts | 1.9 (0.5-4.8) | 0.0 (0.0-1.7) | 3.0 (1.4-5.6) | 0.7 (0.1-2.4) | 2.5 (1.4-4.3) | 0.4 (0.0-1.4) |
| **Any Hands-On SV** | **15.9 (11.3-21.5)** | **2.3 (0.8-5.4)** | **35.1 (29.7-40.8)** | **2.7 (1.2-5.2)** | **27.1 (23.3-31.2)** | **2.5 (1.4-4.3)** |
| **Any Sexual Abuse** | **13.6 (9.3-18.9)** | **2.3 (0.8-5.4)** | **33.8 (28.4-39.4)** | **2.3 (0.9-4.8)** | **25.3 (21.6-29.3)** | **2.3 (1.2-4.1)** |
| Kissing | 8.9 (5.4-13.5) | 1.9 (0.5-4.7) | 21.1 (16.6-26.6) | 1.7 (0.5-3.9) | 16.0 (12.9-19.4) | 1.8 (0.8-3.3) |
| Touching in care | 0.9 (0.1-3.3) | 0.5 (0.0-2.6) | 5.4 (3.1-8.5) | 0.7 (0.1-2.4) | 3.5 (2.1-5.5) | 0.6 (0.1-1.7) |
| Fondling/rubbing | 6.1 (3.3-10.2) | 1.9 (0.5-4.7) | 16.4 (12.4-21.1) | 1.7 (0.5-3.9) | 12.1 (9.4-15.2) | 1.8 (0.8-3.3) |
| Forced undressing | 1.9 (0.5-4.7) | 1.9 (0.5-4.7) | 3.0 (1.4-5.6) | 1.7 (0.5-3.9) | 2.5 (1.4-4.3) | 1.8 (0.8-3.3) |
| **Any Rape** | **3.3 (1.3-6.6)** | **0.0 (0.0-1.7)** | **8.4 (5.5-12.1)** | **1.0 (0.2-2.9)** | **6.2 (4.3-8.7)** | **0.6 (0.1-1.7)** |
| Oral penetration | 0.5 (0.0-2.6) | 0.0 (0.0-1.7) | 1.7 (0.5-3.9) | 0.0 (0.0-1.2) | 1.2 (0.4-2.5) | 0.0 (0.0-0.7) |
| Attempt of oral penetration | 1.4 (0.3-4.0) | 0.0 (0.0-1.7) | 3.3 (1.6-6.1) | 0.3 (0.0-1.8) | 2.5 (1.4-4.3) | 0.2 (0.0-1.1) |
| Vaginal or anal penetration | 0.9 (0.1-3.3) | 0.0 (0.0-1.7) | 4.3 (2.3-7.3) | 0.3 (0.0-1.8) | 2.9 (1.6-4.8) | 0.2 (0.0-1.1) |
| Attempt of vag. or anal penetr. | 0.9 (0.1-3.3) | 0.0 (0.0-1.7) | 2.0 (0.7-4.3) | 0.3 (0.0-1.8) | 1.6 (0.7-3.1) | 0.2 (0.0-1.1) |
| Forcing to penetrate | 0.0 (0.0-1.7) | 0.0 (0.0-1.7) | 0.3 (0.0-1.8) | 0.0 (0.0-1.2) | 0.2 (0.0-1.1) | 0.0 (0.0-0.7) |

Abbreviations: SV = Sexual Violence, CI = Confidence Interval

### 3.2 Coercion strategies

Figure 1 shows the types of coercion used by the assailant for the different types of hands-on lifetime SV. Over one third of the victims indicated that none of the provided response options applied to their situation. For (attempted) rape specifically, (threat of) using physical force was the most commonly identified coercion strategy.

**Figure 1.**
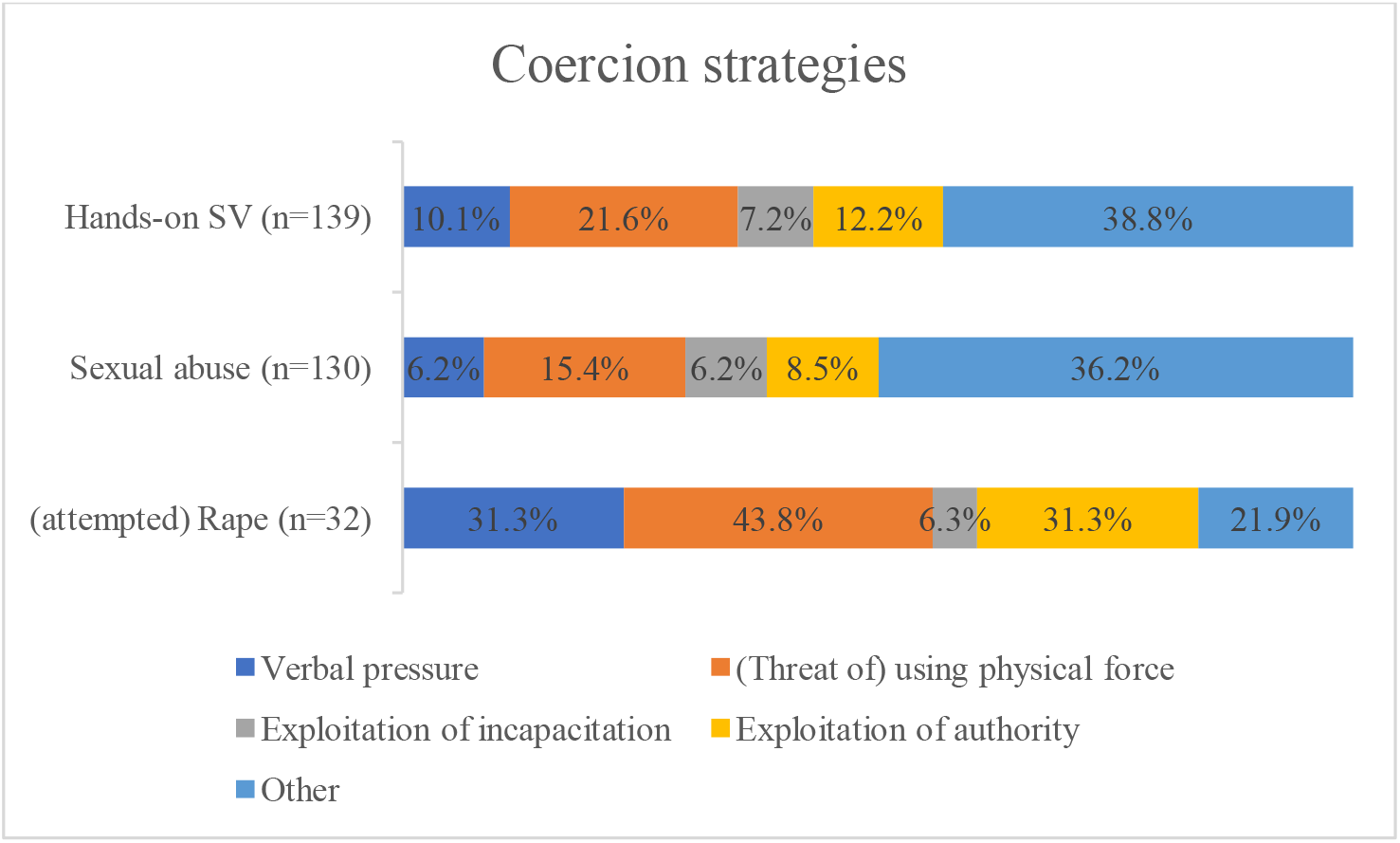
Type of coercion used for hands-on sexual violence, sexual abuse, and (attempted) rape^a^. Note. Respondents could provide multiple answers, unless “Other”= None of the above was selected. Abbreviations: SV= sexual violence

### 3.3 Characteristics of SV victims

Table 2 summarizes the results of the logistic regression analysis. Women were more likely to be sexually victimised in their lifetime, but for the past 12-months we found no difference between women and men regarding sexual victimisation. Participants with two or more lifetime sexual partners experienced more SV in their life compared to participants with fewer than two sexual partners. This difference was not significant in the past 12-months. Regarding sexual orientation, we found that older adults who identified themselves as non-heterosexual experienced significantly more SV in the past 12-months. However, for lifetime SV this difference was not significant.

**Table 2.** Sexual Violence Victimisation Correlates

| Predictors |  | Lifetime SV<br>aOR (95% CI) | Past 12-months SV<br>aOR (95% CI) |
| --- | --- | --- | --- |
| Sex | Female | **3.60 (2.35-5.52) \*** | 1.57 (0.74-3.34) |
|  | Male | Ref | Ref |
| Perceived age | Younger | 1.43 (0.87-2.36) | 0.85 (0.36-2.00) |
|  | Same | Ref | Ref |
|  | Older | 0.85 (0.29-2.46) | / |
| Sexual orientation | Heterosexual | Ref | Ref |
|  | Non-heterosexual | 0.80 (0.38-1.70) | **3.23 (1.17-8.94) \*** |
| Living situation | Community-dwelling | Ref | Ref |
|  | Assisted living | 2.01 (0.78-5.20) | 0.97 (0.20-4.62) |
|  | Nursing home | 0.70 (0.26-1.91) | 0.94 (0.19-4.85) |
| Relationship status | No partner | Ref | Ref |
|  | Not living with partner | 0.96 (0.62-1.49) | 0.51 (0.23-1.14) |
|  | Living with partner | 0.67 (0.30-1.53) | 0.21 (0.03-1.69) |
| Education level | Primary or none | 0.75 (0.44-1.29) | 0.60 (0.24-1.52) |
|  | Secondary | 0.87 (0.55-1.36) | 0.54 (0.25-1.19) |
|  | Higher | Ref | Ref |
| Financial status | Easy | Ref | Ref |
|  | Difficult | 1.02 (0.66-1.60) | 0.67 (0.29-1.55) |
| Care dependency | No | Ref | Ref |
|  | Yes | 1.04 (0.68-1.59) | 0.91 (0.43-1.94) |
| Social support |  | 1.01 (0.99-1.04) | 1.00 (0.96-1.04) |
| Perceived health status | No disability/chronical illness | Ref | Ref |
|  | Disability/chronical illness | 0.96 (0.64-1.45) | 0.84 (0.41-1.74) |
| Sexual initiation<sup>a</sup> | Early (<21 years) | 1.25 (0.84-1.86) | 1.16 (0.56-2.39) |
|  | Late (≥21 years) | Ref | Ref |
| N of lifetime sexual partners<sup>a</sup> | <2 | Ref | Ref |
|  | ≥ 2 | **1.54 (1.01-2.34)\*** | 1.93 (0.92-4.04) |

Abbreviations: SV = Sexual violence, aOR = adjusted odds ratio
\* $p < .05$
<sup>a</sup>Sexual initiation and N of lifetime sexual partners were dichotomised based on the median.

### 3.4 Assailant characteristics

For lifetime SV, 83.6% of assailants were male, 15.0% were female, and in 1.4% of the cases the sex of the assailant was unknown. In the past 12-months, 73.3% of assailants were male, 24.4% were female and in 0.2% of the cases the sex of the assailant was unknown. Mean age of the assailant committing SV in the past 12-months, as estimated by the victim, was 48.9 years (SD 18.9yrs). For both lifetime and past 12-months SV ‘someone unknown’ was most often identified as the assailant, respectively in 41.4% and 44.2% of the cases. More details on the relationship between victim and assailant can be found in Figure 2.

**Figure 2.**
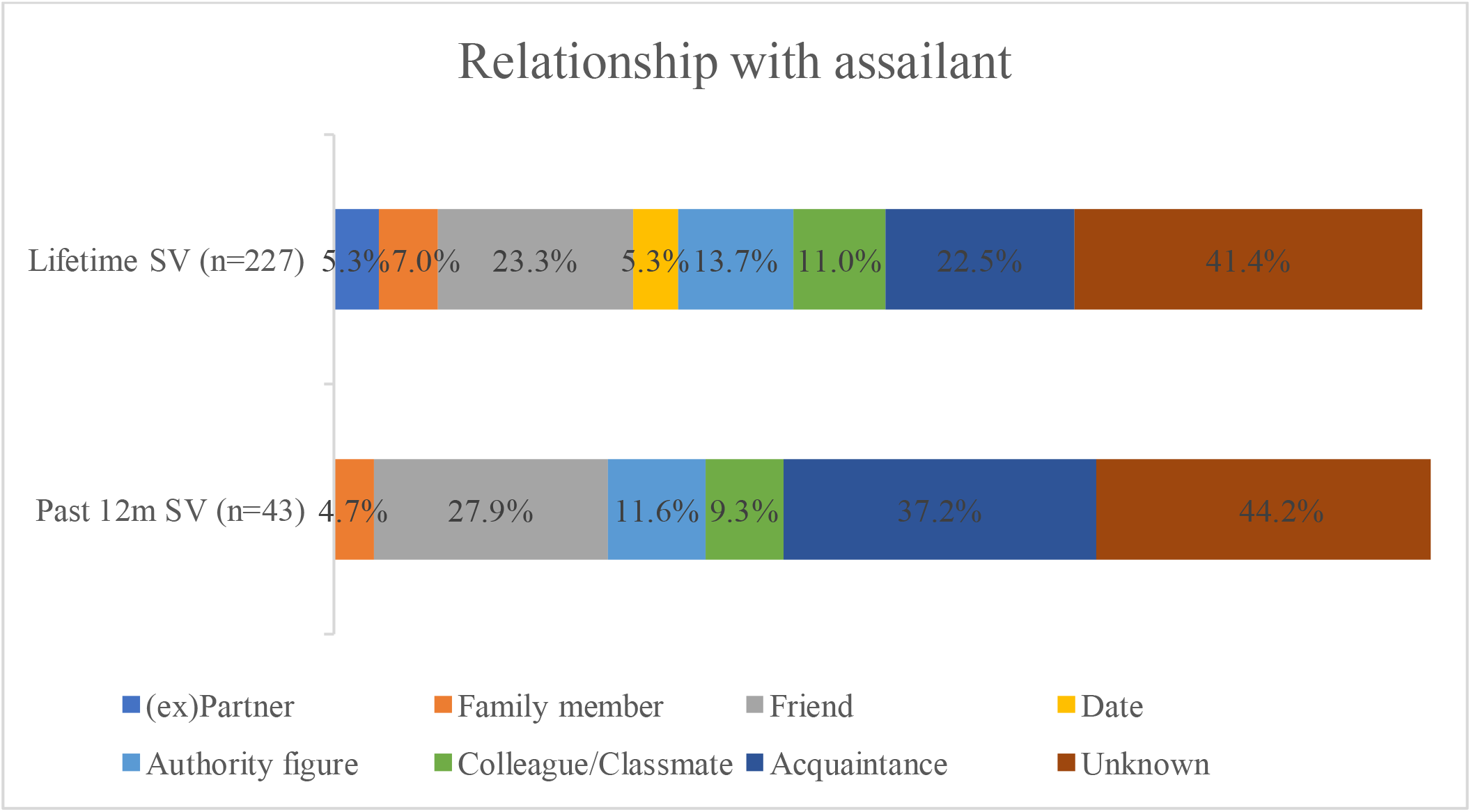
Relationship between victim and assailant of sexual violence, in %. Note. Participants could provide multiple answers. Abbreviations: SV= sexual violence, Past 12m = past 12-months

### 3.5 Framing of sexual violence by victims

Figure 3 summarizes how victims framed SV. In 47.6% of the cases, SV was framed as ‘just something that happened’, in 34.4% as ‘wrong, but not a crime’ and in 23.3% as a crime. Concerning rape, we found that in 28.1% of cases victims framed it as ‘just something that happened’, in 28.1% as ‘wrong, but not a crime’, and in 43.8% as a crime.

**Figure 3.**
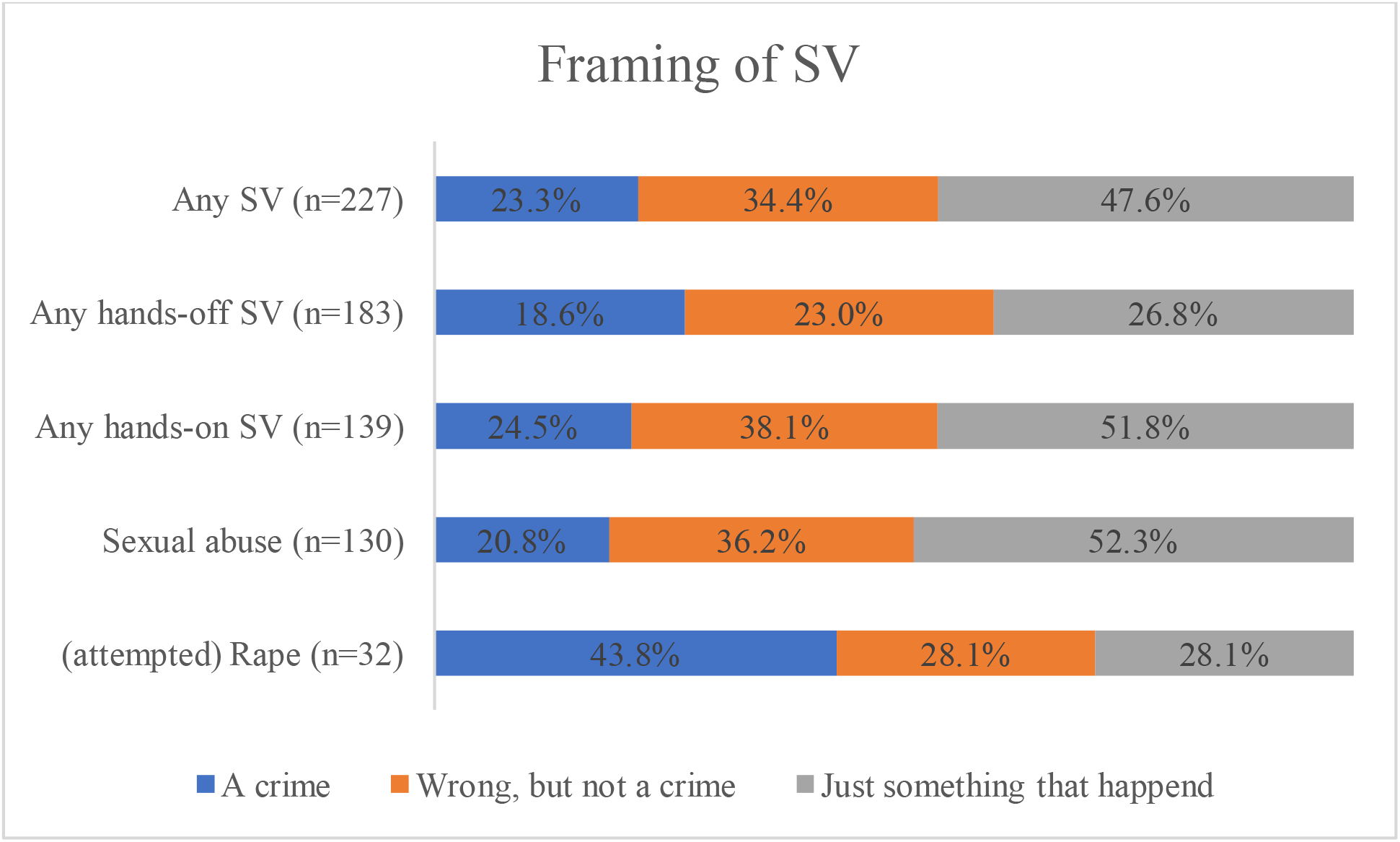
Framing of sexual violence by victims, in %. Note. For hands-off SV only the incident with the most impact on the victim was included in the analysis. For hands-on SV all incidents were included and grouped into sexual abuse, (attempted) rape and hands-on SV. If victims indicated a different framing for different incidents, they are included as separate answers and so the total % is >100%. Abbreviations: SV= sexual violence

## 4. Discussion

In this paper we present a Belgian prevalence study on sexual victimisation in older adults. We conducted 513 interviews with people aged 70 years and older across Belgium.

Our results show that lifetime exposure to SV is highly prevalent among older adults in Belgium. Over 44% of participants were sexually victimized during their lifetime. Despite the assumption that older adults are at low risk for sexual victimisation [7], in our study, one in 12 older adults experienced at least one form of SV in the past 12-months. Our numbers appear higher compared to previous European studies in community-dwelling older adults in which the estimated lifetime SV prevalence was 6.3% and the past 12-months prevalence rates varied between 0 and 3.1% [9, 10]. This difference could be explained by several methodological choices. First, we studied SV in older adults from a different perspective compared to previous studies that researched SV based on criminal cases [12, 13] or as a form of elder abuse and neglect, domestic violence and intimate partner violence [9, 11]. Hence, they restricted the relation between victim and assailant to a confidant, a household member or an intimate partner respectively while our research shows that assailants are also unknown. Moreover, previous research only included forms of hands-on SV (sexual abuse with physical contact and (attempted) rape). Applying the broad WHO definition of SV, we included both hands-off and hands-on SV regardless of the relation between victim and assailant, leading to increased lifetime and past 12-months SV prevalence numbers. Second, the use of the BSQ made it easier for participants to remember and engage with the situations presented. Furthermore, BSQ leaves less room for interpretation, stigma or labelling which makes it possible for people who do not identify as a victim to indicate their SV experiences, leading to more valid estimates [26].

However, compared to an online study in the Belgian population aged 16 to 69 years using the same questionnaire, we found lower lifetime and past 12-months prevalence rates [34]. This decreased SV reporting with increasing age may adequately represent lower sexual victimisation rates in older adults or may be explained by several factors, such as reduced recall in general [35], reduced recall of negative events [36, 37], or higher mortality among people with a SV history [38, 39]. Moreover, older adults might have a different perception of SV than younger generations. In our study, in 47.6% of SV cases and in 28.1% of rape cases, victims perceived it as ‘just something that happened’. Previous studies found that generational specificities surrounding sexuality and SV such as legal definitions and perceptions of SV, influenced disclosure rates [13, 40]. Furthermore, society’s attitudes regarding sexuality have become more permissive, and the definition of sexual consent has been narrowed [14]. For example, until the end of the 20^th^ century being married implied consent to sexual intercourse, whereas today spousal rape is considered a criminal offence [15]. In our study, in only 5.3% of lifetime SV cases, the (ex)partner was identified as assailant, which is much less compared to studies in younger populations in which over 25% of women indicated being sexually victimised by their (ex) partner [41].

Finally, because of the image of ‘the asexual older adult ‘[17], older adults might not identify themselves as a victim of SV [18, 19]. In previous studies on elder abuse and neglect, older adults did not acknowledge SV as a possible form of abuse [42]. To a certain extent, we have pre-emptively addressed this by adopting BSQ to measure sexual victimisation. Nevertheless, such beliefs may have inadvertently influenced SV disclosure in our study.

In addition to measuring SV prevalence, our study aimed to provide an analysis of SV correlates in older adults. Correlates generally linked to elder abuse and neglect such as poor (perceived) health status, care dependency, low social support, and financial strain [43-48], were not associated with sexual victimisation in our sample. Being female and having a greater number of lifetime sexual partners were associated with lifetime sexual victimisation, which is in line with previous research on SV in younger populations [28, 49]. For past 12-months SV, we could not identify a difference between men and women. Previous research showed inconclusive results. Although some studies described older women as being more prone to SV [50, 51], others showed that women and men were equally at risk [52, 53]. In our sample, being non-heterosexual was correlated to past 12-months SV. Previous research has linked LGBT+ status, often intertwined with other factors such as disability and poverty, to intimate partner violence among older adults [54], but for SV this has not been reported before. However, our results have to be interpreted with caution as a possible difficulty of several participants to understand the different terms defining sexual orientation, could lead to an overestimation of non-heterosexual people in our sample. Furthermore, our results confirm previous findings that assailants of SV in older adults tend to be younger than the victim [50].

Regarding coercion strategies, we found that the (threat of) using physical force was the most common coercion used for any type of rape. For any type of sexual abuse, over one third of the participants indicated that none of the mentioned types of coercion were used. Previous studies showed inconclusive results regarding coercion strategies. Although some studies reported physical force was more often used on older SV victims compared to younger victims, most studies did not report significant differences between younger and older victims regarding use of physical force as a coercion strategy [12]. Because our findings are similar to the coercion strategies identified by younger victims in Belgium [34], we assume that the coercion strategies used on older adults are similar to the ones used on younger victims and not as violent as believed [12].

An important limitation of our study was that the target sample size of 845 interviews could not be reached due to the COVID-19 pandemic and associated lockdown measures. However, the current sample size of 513 interviews allowed us to report on prevalence rates within four percent of the estimated value. Furthermore, due to the absence of a standardised measure for sexual neglect, we narrowed it down to “touching in care” which is an incomplete representation of the definition [25] and supposed reality. Nevertheless, this study is, to our knowledge, the first of its kind to measure the prevalence, correlates, assailant characteristics and framing of SV in older adults. It can be regarded as an important step towards a better understanding of the magnitude, nature and impact of SV in older adults. Responding to the call of Bows [12] to consider SV as a particular form of violence in old age and study it independently from other forms of elder abuse and neglect and domestic violence, this study brings a new perspective on SV in older adults. For future studies, we encourage the development of measurement tools for sexual neglect in order to incorporate this form of SV as well. Based on our findings we reinforce previous recommendations for policy makers to recognise older adults as a risk group for sexual victimisation [12]. Furthermore, our study showed that assessing SV in older adults is possible without offending them [31]. Professionals urgently need capacity building to better detect signs, prevent, mitigate and respond to SV in old age. Finally, sensitisation of society in general is essential, emphasizing the prevention of SV against older adults.

## 5. Conclusion

Sexual victimisation appears to be common in older adults in Belgium. Over 44% experienced SV in their lifetime and one in 12 in the past 12-months. Being female and having had a greater number of lifetime sexual partners were linked to lifetime SV, a non-heterosexual sexual orientation to past 12-months victimisation. Correlates generally linked to elder abuse and neglect did not seem to be linked with SV. Our findings highlight the importance of recognising older adults as a risk group for sexual victimisation and to study SV independently from other forms of violence in old age. In order to detect signs, prevent, mitigate and respond to SV in older adults, sensitisation of society and capacity building of professionals is needed.

## Data Availability

The data that support the findings of this study are available from the corresponding author, A.N., upon reasonable request.

## Acknowledgements

Anne Nobels and Adina Cismaru Inescu contributed equally to this work and are therefore to be regarded as joint first authors of this article. Ines Keygnaert and Christophe Vandeviver contributed equally to this work and are therefore to be regarded as joint last authors of this article.

The authors would like to thank Lotte De Schrijver and Joke Depraetere for their input during the questionnaire development. Also, we want to thank our interviewers for their time and dedication to our study, and Dr. Howard Ryland for his help with the language editing. Finally, we want to thank all older adults who participated in our study for sharing their stories with us.

## Appendices Appendix 1. Detailed outcome measurements sexual victimisation

### Hands-off sexual victimisation (no physical contact)

- *Sexual staring:* Someone stared at me in a sexual way or looked at my intimate body parts (e.g., breasts, vagina, penis, anus) when I didn’t want it to happen.
- *Sexual innuendo:* Someone made teasing comments of a sexual nature about my body or appearance even though I didn’t want it to happen.
- *Showing sexual images:* Someone showed me sexual or obscene materials such as pictures, videos, directly or over the internet (including email, social networks and chat platforms) even though I didn’t want to look at them. This does not include mass mailings or spam.
- *Sexual calls or texts:* Someone made unwelcome sexual or obscene phone calls or texts to me.
- *Voyeurism:* I caught someone watching me, taking photos or filming me when I didn’t want it to happen while I was undressing, nude or having sex.
- *Distribution of sexual images:* Someone distributed naked pictures or videos of me directly or over the internet (including email, social networks and chat platforms) when I didn’t want it to happen.
- *Exhibitionism:* Someone showed their intimate body parts (e.g., breasts, vagina, penis, anus) to me in a sexual way and/or masturbated in front of me when I didn’t want to see it.
- *Forcing to show intimate body parts:* Someone made me show my intimate body parts (e.g., breasts, vagina, penis, anus) online or face-to-face when I didn’t want to do it.

### Hands-on sexual victimisation

#### Sexual abuse (physical contact but no penetration)

- *Kissing:* Someone kissed me against my will.
- *Touching in care:* Someone touched my intimate body parts (e.g., breasts, vagina, penis, anus) during care against my will.
- *Fondling/rubbing:* Someone fondled or rubbed up against my intimate body parts (e.g., breasts, vagina, penis, anus) against my will.
- *Forced undressing:* Someone removed (some of) my clothes against my will.

#### Rape and attempted rape (physical contact with attempted or completed penetration)

- *Oral penetration:* Someone had oral sex with me or made me give oral sex against my will.
- *Attempt of oral penetration:* Someone tried, but did not succeed, to have oral sex with me or tried to make me give oral sex against my will.
- *Vaginal or anal penetration:* Someone put their penis, finger(s) or object(s) into my vagina or anus against my will.
- *Attempt of vaginal or anal penetration:* Someone tried, but did not succeed to put their penis, finger(s) or object(s) into my vagina or anus against my will.
- *Forcing to penetrate:* Someone made me put my penis, finger(s) or object(s) into their (or someone’s) vagina or anus against my will.

## Appendix 2. Sociodemographic characteristics of the study population (n=513)

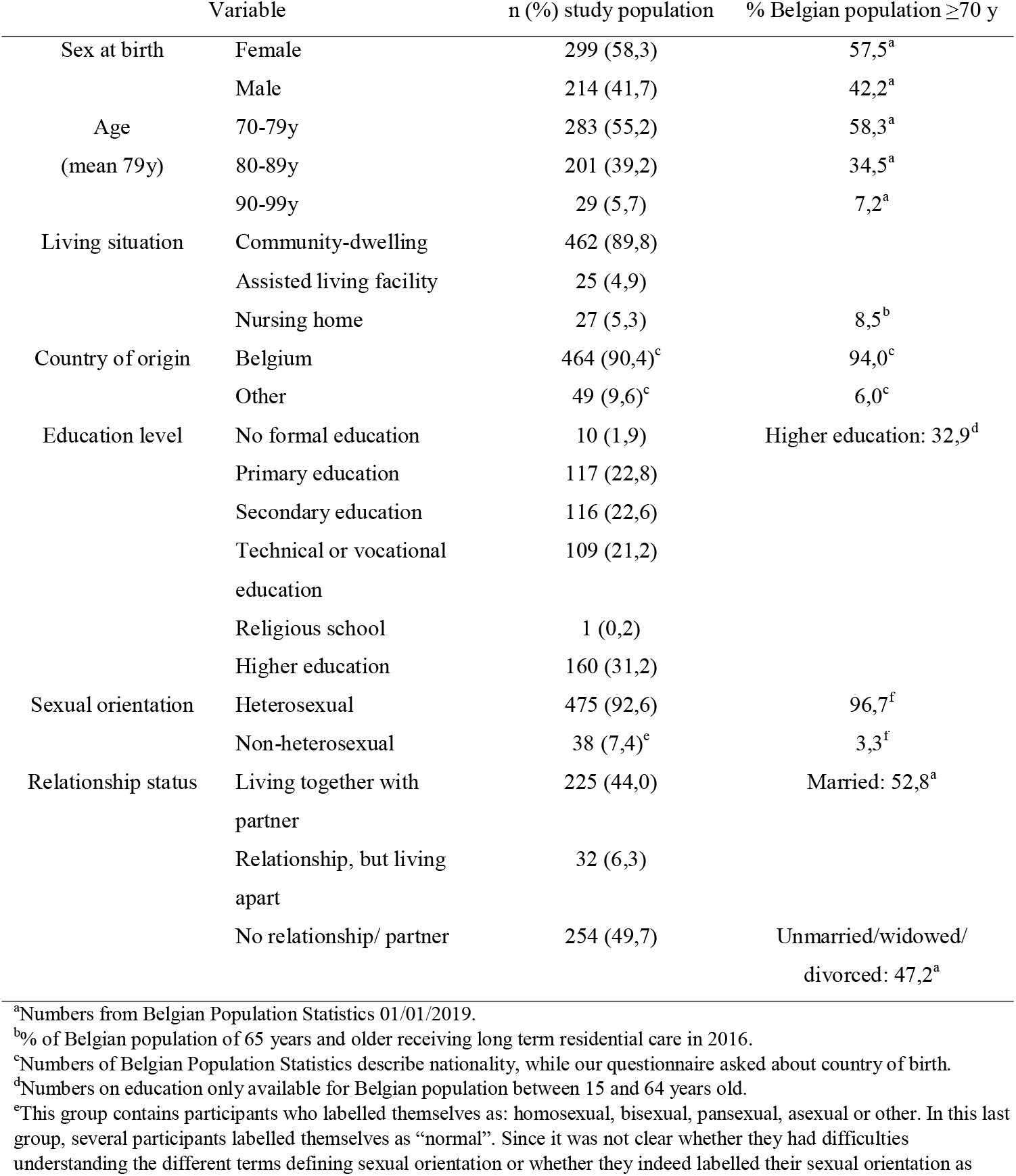
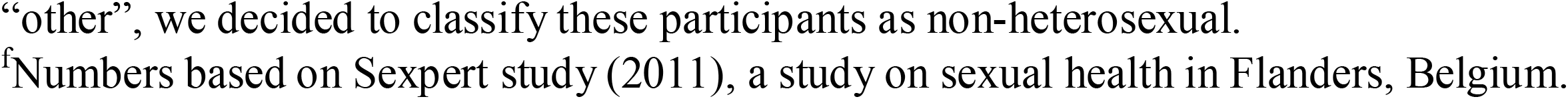

|  | Variable | n (%) study population | % Belgian population ≥70 y |
| --- | --- | --- | --- |
| Sex at birth | Female | 299 (58,3) | 57,5<sup>a</sup> |
|  | Male | 214 (41,7) | 42,2<sup>a</sup> |
| Age<br>(mean 79y) | 70-79y | 283 (55,2) | 58,3<sup>a</sup> |
|  | 80-89y | 201 (39,2) | 34,5<sup>a</sup> |
|  | 90-99y | 29 (5,7) | 7,2<sup>a</sup> |
| Living situation | Community-dwelling | 462 (89,8) |  |
|  | Assisted living facility | 25 (4,9) |  |
|  | Nursing home | 27 (5,3) | 8,5<sup>b</sup> |
| Country of origin | Belgium | 464 (90,4)<sup>c</sup> | 94,0<sup>c</sup> |
|  | Other | 49 (9,6)<sup>c</sup> | 6,0<sup>c</sup> |
| Education level | No formal education | 10 (1,9) | Higher education: 32,9<sup>d</sup> |
|  | Primary education | 117 (22,8) |  |
|  | Secondary education | 116 (22,6) |  |
|  | Technical or vocational education | 109 (21,2) |  |
|  | Religious school | 1 (0,2) |  |
|  | Higher education | 160 (31,2) |  |
| Sexual orientation | Heterosexual | 475 (92,6) | 96,7<sup>f</sup> |
|  | Non-heterosexual | 38 (7,4)<sup>e</sup> | 3,3<sup>f</sup> |
| Relationship status | Living together with partner | 225 (44,0) | Married: 52,8<sup>a</sup> |
|  | Relationship, but living apart | 32 (6,3) |  |
|  | No relationship/ partner | 254 (49,7) | Unmarried/widowed/divorced: 47,2<sup>a</sup> |

<sup>a</sup>Numbers from Belgian Population Statistics 01/01/2019.
<sup>b</sup>% of Belgian population of 65 years and older receiving long term residential care in 2016.
<sup>c</sup>Numbers of Belgian Population Statistics describe nationality, while our questionnaire asked about country of birth.
<sup>d</sup>Numbers on education only available for Belgian population between 15 and 64 years old.
<sup>e</sup>This group contains participants who labelled themselves as: homosexual, bisexual, pansexual, asexual or other. In this last group, several participants labelled themselves as "normal". Since it was not clear whether they had difficulties understanding the different terms defining sexual orientation or whether they indeed labelled their sexual orientation as
“other”, we decided to classify these participants as non-heterosexual.
<sup>f</sup>Numbers based on Sexpert study (2011), a study on sexual health in Flanders, Belgium.

## Notes

Declaration of interest: None.

### Competing Interest Statement

The authors have declared no competing interest.

### Funding Statement

This research was predominantly supported by the Belgian Federal Science Policy Belgian Research
Action through the Interdisciplinary Networks funding scheme, grant number BR/175/A5/UN-MENAMAIS.
CVʹs contribution was supported in part by the Research Foundation Flanders (FWO) Postdoctoral Fellowship funding scheme, grant numbers 12C0616N, 12C0619N.

### Author Declarations

Ethical committee of Ghent University/University Hospital (Belgian registration number B670201837542)

